# Surface and air contamination with SARS-CoV-2 from hospitalized COVID-19 patients in Toronto, Canada

**DOI:** 10.1101/2021.05.17.21257122

**Authors:** Jonathon D. Kotwa, Alainna J. Jamal, Hamza Mbareche, Lily Yip, Patryk Aftanas, Shiva Barati, Natalie G. Bell, Elizabeth Bryce, Eric Coomes, Gloria Crowl, Caroline Duchaine, Amna Faheem, Lubna Farooqi, Ryan Hiebert, Kevin Katz, Saman Khan, Robert Kozak, Angel X. Li, Henna P. Mistry, Mohammad Mozafarihashjin, Jalees A. Nasir, Kuganya Nirmalarajah, Emily M. Panousis, Aimee Paterson, Simon Plenderleith, Jeff Powis, Karren Prost, Renée Schryer, Maureen Taylor, Marc Veillette, Titus Wong, Xi Zoe Zhong, Andrew G. McArthur, Allison J. McGeer, Samira Mubareka

## Abstract

**Background:** The aim of this prospective cohort study was to determine the burden of SARS-CoV-2 in air and on surfaces in rooms of patients hospitalized with COVID-19, and to identify patient characteristics associated with SARS-CoV-2 environmental contamination.

**Methods:** Nasopharyngeal swabs, surface, and air samples were collected from the rooms of 78 inpatients with COVID-19 at six acute care hospitals in Toronto from March to May 2020. Samples were tested for SARS-CoV-2 viral RNA and cultured to determine potential infectivity. Whole viral genomes were sequenced from nasopharyngeal and surface samples. Association between patient factors and detection of SARS-CoV-2 RNA in surface samples were investigated using a mixed-effects logistic regression model.

**Findings:** SARS-CoV-2 RNA was detected from surfaces (125/474 samples; 42/78 patients) and air (3/146 samples; 3/45 patients) in COVID-19 patient rooms; 17% (6/36) of surface samples from three patients yielded viable virus. Viral sequences from nasopharyngeal and surface samples clustered by patient.

Multivariable analysis indicated hypoxia at admission, a PCR-positive nasopharyngeal swab with a cycle threshold of ≤30 on or after surface sampling date, higher Charlson co-morbidity score, and shorter time from onset of illness to sample date were significantly associated with detection of SARS-CoV-2 RNA in surface samples.

**Interpretation:** The infrequent recovery of infectious SARS-CoV-2 virus from the environment suggests that the risk to healthcare workers from air and near-patient surfaces in acute care hospital wards is likely limited. Surface contamination was greater when patients were earlier in their course of illness and in those with hypoxia, multiple co-morbidities, and higher SARS-CoV-2 RNA concentration in NP swabs. Our results suggest that air and surfaces may pose limited risk a few days after admission to acute care hospitals.

## Introduction

Severe acute respiratory syndrome coronavirus 2 (SARS-CoV-2) emerged in December 2019 causing the coronavirus disease 2019 (COVID-19) pandemic^1^ and many hospital outbreaks of COVID-19.^2^ Understanding the role of surface and air (environmental) contamination in the transmission of SARS-CoV-2 is essential to ensuring the prevention of transmission of SARS-CoV-2 between patients and to healthcare workers in acute care hospitals.

SARS-CoV-2 RNA has been detected from surfaces and air in hospitals.^3–12^ However, a minority of studies have attempted to culture virus.^13,14^ This limits our understanding of exposure and transmission risk.

SARS-CoV-2 whole-genome sequencing has been used to determine the extent of community transmission and inform public health intervention.^15^ While whole-genome sequences are generally obtained through upper respiratory samples such as nasopharyngeal (NP) swabs,^16–18^ applying a genomics approach to environmental samples for SARS-CoV-2 may confirm sources of environmental contamination and potentially identify transmission bottlenecks and determinants of environmental persistence.

This study aimed to determine the burden of SARS-CoV-2 in the air and on surfaces in hospital rooms of acutely ill inpatients with COVID-19 in Toronto, Ontario, Canada. We also compared SARS-CoV-2 whole-genome viral sequences from patient nasopharyngeal swabs and surfaces in their rooms and determined the association between patient factors and detection of SARS-CoV-2 from environmental samples.

## Methods

### Study population

The Toronto Invasive Bacterial Diseases Network (TIBDN) performs population-based surveillance for infectious diseases in metropolitan Toronto and the regional Municipality of Peel, south-central Ontario, Canada (population 4·2 million in 2016). TIBDN clinical microbiology laboratories report clinical specimens yielding SARS-CoV-2 to TIBDN’s central study office. At six TIBDN hospitals, consecutive inpatients with laboratory confirmed COVID-19 identified between March and May 2020 were eligible for this study. Research ethics approval was granted by all participating The Toronto Invasive Bacterial Diseases Network hospitals (Sunnybrook’s Research Ethics Board, The Mount Sinai Hospital Research Ethics Board, Toronto East Health Network Research Ethics Board, Osler’s Research Ethics Board, and Scarborough Health Network’s Research Ethics Board). Informed consent from all patients were obtained. Findings were reported in accordance with the Strengthening the Reporting of Observational studies in Epidemiology (STROBE) guidelines for reporting observational studies.^19^

### Data and specimen collection

Demographic, clinical, and COVID-19 risk factor data were collected by participant interview and chart review. Study staff obtained NP swabs from patients at enrollment and every three days until refusal, hospital discharge, or death.^20^ A set of surface samples was collected at enrollment and every three days, including: 1) bathroom doorknob, 2) phone (all surfaces of the patient’s phone and room phone), 3) overbed table and chair (pooled), 4) bed (bed rail and pillow) and light switch or pullcord in patient’s bedspace (pooled), and 5) toilet and sink faucet handles (pooled) (Supplementary Figure 1). Surface samples were collected by thoroughly wiping each surface type using the rough side of a dry 6 cm x 6 cm Swiffer cloth (Swiffer®, Procter & Gamble, Toronto, Canada). Nasopharyngeal swabs and Swiffer cloths were immediately placed into universal transport medium (UTM; Copan Diagnostics, Murrietta, CA).

During the study period, four bioaerosol samplers were used for sampling the first 45 patients enrolled that were not intubated. For each patient, one to two different bioaerosol samplers were used in each run. Using an air sampling pump (GilAir Plus Personal Air Sampling Pump, Sensidyne, St. Petersburg, FA), air samples were obtained using the 1 μm pore size, 37 mm polytetrafluoroethylene (PTFE) membrane filters (SKC Inc, Eighty Four, PA), the 37 mm three-piece cassette with 0·8 μm polycarbonate (PC) filter (Zefon International, Ocala, FA), and 25 mm gelatin membrane filters (SKC Inc, Eighty Four, PA). Prior to sampling, the pumps were calibrated to a flow rate of 3·5 L/min using the corresponding filter used for sampling that day (Gilibrator 3, Standard Flow Dry Cell Calibrator, Sensidyne, St. Petersburg, FA). In the patient rooms, samplers were placed at 1 m and 2 m from the patient at the level of the bed and samples were collected over a 2 h period. All filters were placed in coolers at the end of the sampling period for transport and processed immediately. Air samples were also collected using the NIOSH two-stage cyclone bioaerosol sampler (National Institute for Occupational Safety and Health, Morgantown, WV).

The NIOSH cyclone bioaerosol sampler is comprised of stages collecting larger particles (>4 μm) in the first stage into 15 mL conical tubes, smaller particles (1–4 μm) in the second stage into 1·5 mL conical tubes, and particles <1 μm onto a PTFE filter. The NIOSH cyclone samplers were assembled in the laboratory in a biosafety cabinet and calibrated to a flow rate of 3.5 L/min (BIOS DC-1 DryCal flow calibrator, SKC Inc, Eighty-Four, PA). In the patient rooms, the sampler was placed 1m from the patient and sampling occurred over a 2 h period.

### Laboratory procedures

All samples were processed at Sunnybrook Research Institute on the day of collection. Nasopharyngeal swabs and environmental samples were vortexed for 20 s before aliquoting and storage at -80°C. PTFE, PC, and gelatin membrane filters were placed in 3 ml transport media before being vortexed for 20s, followed by aliquoting and storage at -80°C. For the NIOSH cyclone bioaerosol sampler, 1 mL of transport media was added to the first stage, 500 μL to the second stage, and 3 mL of transport media were aliquoted onto the PTFE filter. Samples were vortexed for 20 s before aliquoting and storage at - 80°C. RNA extractions were performed using QIAmp viral RNA mini kit (Qiagen, https://www.qiagen.com) according to manufacturer’s instructions; samples were eluted into 40 uL. Reverse-transcriptase polymerase chain reaction (RT-PCR) reactions were performed using the Luna Universal Probe One-Step RT-qPCR Kit (New England BioLabs Inc, https://www.international.neb.com). Two separate gene targets were used for detection of SARS-CoV-2, the 5’ untranslated region (UTR) and the envelope (E) gene, with human RNaseP as an internal control.^21^ The cycling conditions were: 1 cycle of denaturation at 60 °C for 10 min then 95 °C for 2 min followed by 44 amplification cycles of 95°C for 10 s and 60°C for 15 s. Rotor-Gene Q software (Qiagen, https://www.qiagen.com) was used to determine cycle thresholds (Ct) and samples with Cts <40 in both UTR and E genes were considered positive. Correlation analysis indicates almost perfect correlation between Ct values for the UTR and E gene (0·99). We therefore present Ct values for the UTR gene target within the text; the Ct value results for both gene targets are summarized in Figure 1. See Supplementary Methods for details on genome sequencing and analysis.

**Figure 1.**
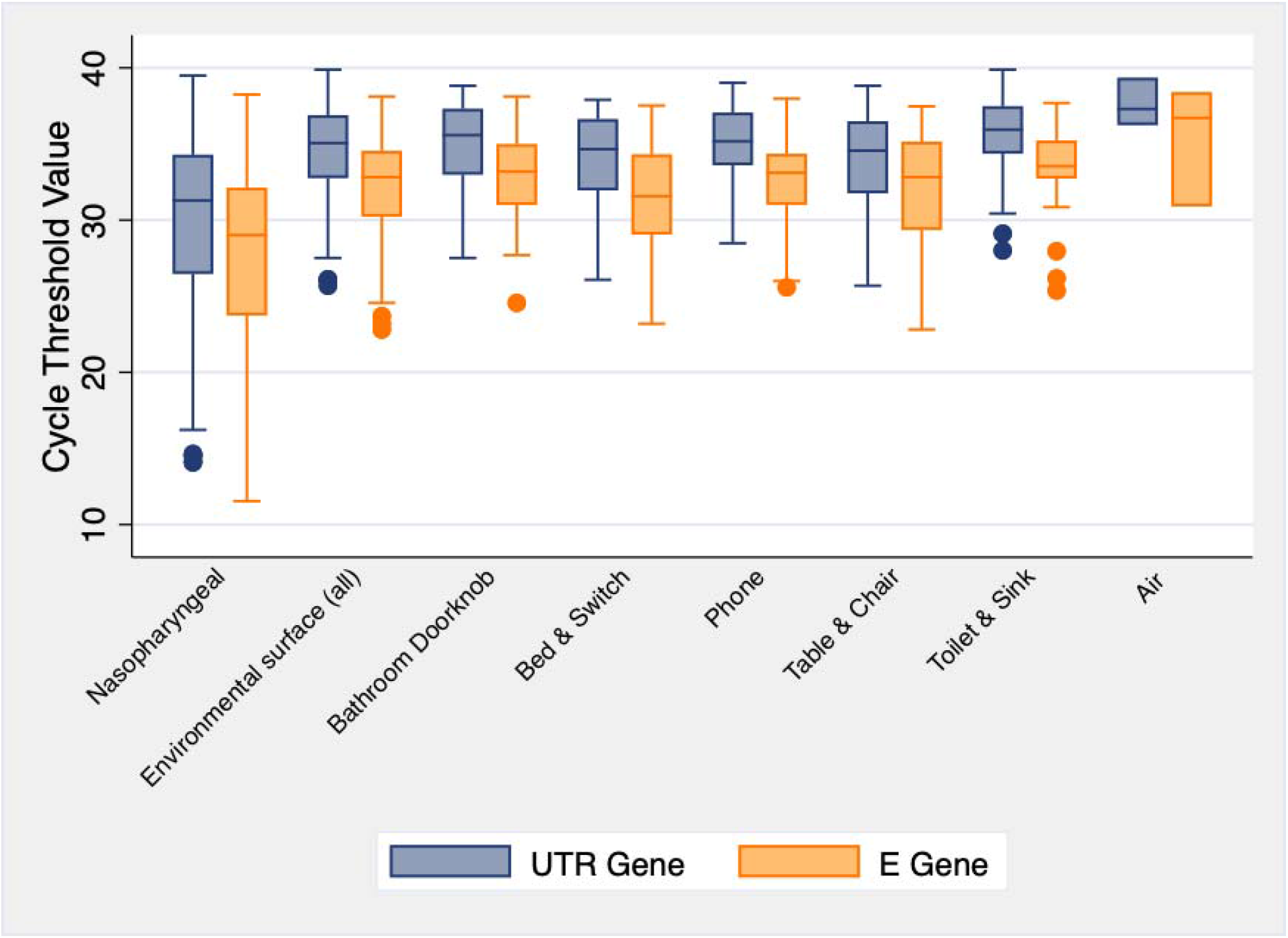
Boxplot summary of the cycle threshold values for the UTR gene (blue) and E gene (green) targets from the SARS-CoV-2 PCR analysis for each sample type investigated for 78 COVID-19 positive patients in Toronto, Canada. Notably, air sampling pumps were calibrated to a flow rate of 3·5 L/min for 2 h; each air sample represents 420 L of air.

### Virus isolation

Virus isolation was attempted on PCR-positive NP swabs and air samples, and PCR-positive environmental surface samples with a Ct of <34·0 in containment level 3 at the University of Toronto.

Vero E6 cells were seeded at a concentration of 3×10^5^ cells/well in a six well-plate. The next day, 500 uL of sample containing 16 ug/mL TPCK-treated trypsin (New England BioLabs Inc, https://www.international.neb.com), 2X Pen/Strep and 2X antibiotic-antimycotic (Wisent, https://www.wisentbioproducts.com/en/) were used to inoculate cells. Plates were returned to a 37 °C, 5% CO_2_ incubator for 1 h and rocked every 15 minutes. After 1 h, the inoculum was removed and replaced with DMEM containing 2% FBS, 6 ug/mL TPCK-treated trypsin, 2X Pen/Strep, and 2X antibiotic-antimycotic. Cells were observed daily under a light microscope for cytopathic effect (CPE) for 5 days post infection. Cell cultures not showing any CPE were blind passaged onto fresh Vero cells and observed for a further 5 days. The RT-PCR assay described above was used to confirm SARS-CoV-2 isolation from supernatant.

### Statistical analysis

All analyses were conducted using Stata/SE 15·1 (StataCorp, College Station, Texas, USA; http://www.stata.com). Descriptive statistics were used for patient characteristics, PCR results, and culture results. To explore putative associations with SARS-CoV-2 PCR-positive environmental surface samples we reviewed the literature and surveyed Canadian COVID-19 researchers to identify factors of interest which might be associated with environmental contamination. The following variables were investigated: age, sex, Charlson comorbidity index,^22^ smoking history, Clinical Frailty Score,^23^ presence or absence of symptoms from onset to 24 hours post admission (cough, fever, diarrhea, delirium/confusion), hypoxia at admission (defined as oxygen saturation < 92%), admission to intensive care unit (ICU) at time of sample collection, use of exogenous oxygen during stay, prone position, receiving steroids for treatment on day of sampling, room type (regular private room or negative pressure room), and the presence or absence of a PCR-positive NP swab on or after environmental sampling date (PCR-positive NP swabs were further categorized to Ct>30 and Ct≤30); the sampling date refers to the date the sample was taken. If use of exogenous oxygen during stay was significant, oxygen delivery methods (intubation, facemask/nasal prong, high flow) were included to investigate individual oxygen requirements. Since samples were taken serially from each patient over the course of this study, we included onset of illness to sample date as a fixed-effect control to account for temporal variability. See Supplementary Table 2 for further variable details. The outcome of interest was SARS-CoV-2 PCR-positive environmental surface samples.

A causal diagram was constructed to examine possible confounding and intervening relationships among exploratory variables relative to a SARS-CoV-2 PCR-positive environmental surface sample. Mixed-effects logistic regression models with a random intercept for unique patient identification to account for clustering were constructed using backwards elimination. Variables that were significant, potential confounders, part of a significant interaction term, or a control variable (i.e. onset of illness to sample date) were included in the final model.

Pearson and deviance residuals were explored for outlying observations. Model fit was assessed by determining if the best linear unbiased predictors (BLUPs) met the assumptions of normality and homogeneity of variance.

### Role of the funding source

The funders had no role in study design, data collection, data analysis, data interpretation, or manuscript preparation.

## Results

### Study population and samples collected

There were 78 inpatients with COVID-19 who consented to participate. All were confirmed to have COVID-19 with a positive nasal, mid-turbinate, or NP swab tested in a licensed diagnostic laboratory in Toronto prior to enrollment; diagnostic samples used for initial COVID-19 confirmation were not included in the present study. Patients were de-identified and randomly assigned a number from 1-78.

The median age of participants was 67 years (interquartile range [IQR] 53–79). The median duration between onset of illness and admission was five days (IQR 2–8). Patient characteristics are shown in Table 1. Numbers of additional NP swabs, surface samples, and air samples collected are shown in Table 2; a detailed breakdown of samples collected is shown in Supplementary Table 1.

**Table 1.**
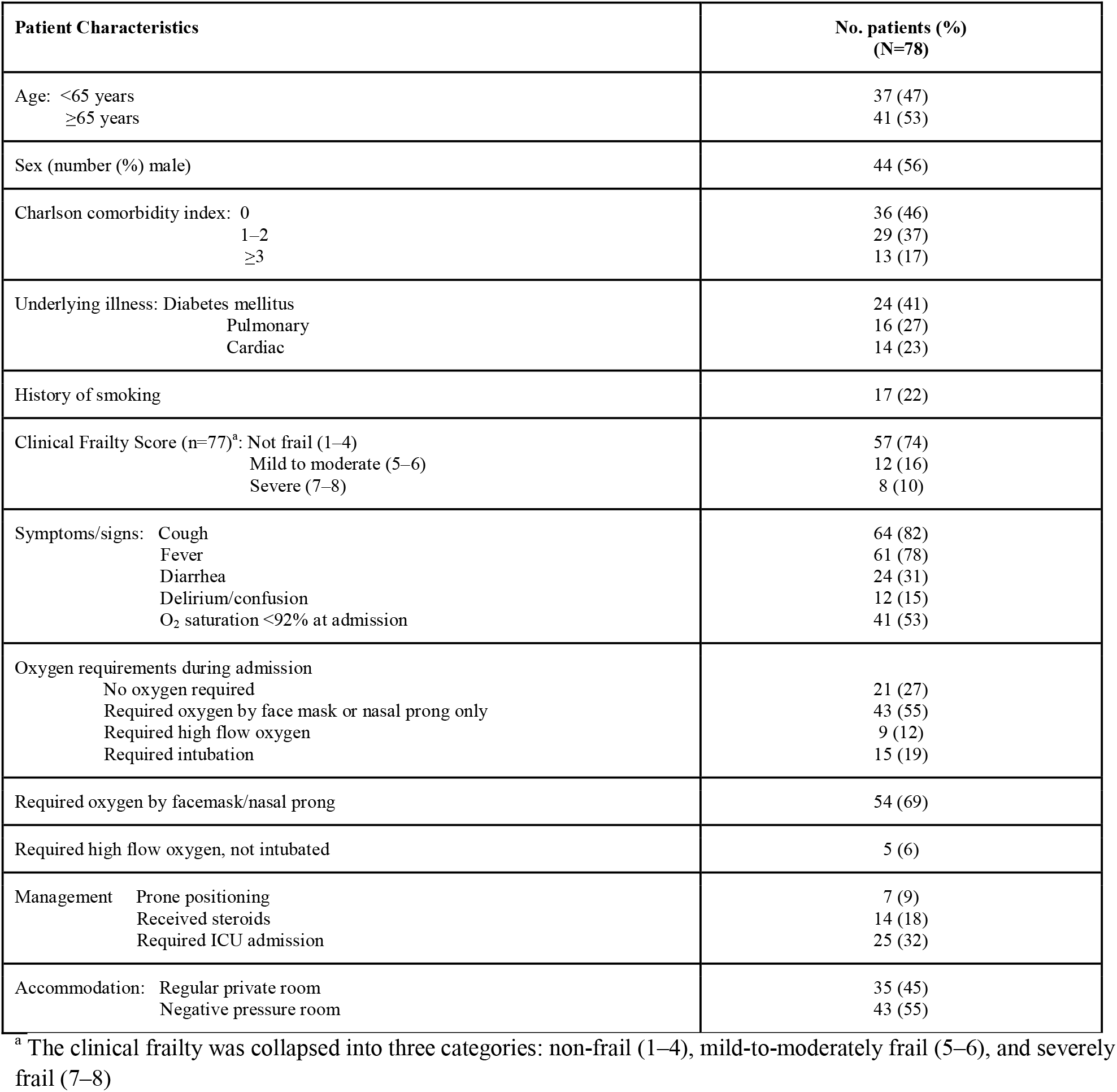
Patient demographics and clinical characteristics for 78 hospitalized patients with COVID-19 in Toronto, Canada.

**Table 2.** Summary of sample types collected and results of PCR testing and cell culture for SARS-CoV-2 in 78 hospitalized patients with COVID-19 in Toronto, Canada.

| Sample type | PCR |  | Culture<sup>b</sup> |  |
| --- | --- | --- | --- | --- |
| | No. positive samples/total (%) | No. patients with $\geq 1$ sample positive/total (%)<sup>a</sup> | No. positive samples/total cultured (%) | No. patient with $\geq 1$ sample positive/total (%) |
| Nasopharyngeal swab | 172/219 (79) | 74/78 (95) | 30/110 (27) | 21/65 (32) |
| Environmental surface (all) | 125/474 (26) | 42/78 (54) | 6/36 (17) | 3/16 (19) |
| Bathroom door | 12/88 (14) | 12/69 (17) | 1/4 (25) | 1/4 (25) |
| Bed and switch (pooled) | 39/102 (38) | 33/78 (42) | 2/13 (15) | 2/11 (18) |
| Phone | 24/88 (27) | 21/70 (30) | 1/5 (20) | 1/4 (25) |
| Table and chair (pooled) | 29/95 (31) | 24/74 (32) | 1/10 (10) | 1/9 (11) |
| Toilet and sink (pooled) | 21/101 (21) | 20/74 (27) | 1/4 (25) | 1/3 (33) |
| Air<sup>c</sup> | 3/146 (2) | 3/45 (7) | 0/3 (0) | 0/3 (0) |
| 1 m from head of patient | 3/101 (3) | 3/45 (7) | 0/3 (0) | 0/3 (0) |
| 2 m from head of patient | 0/45 (0) | 0/45 (0) | .. | .. |

<sup>a</sup> Total number of patients for each category differ as samples were taken every three days post enrollment until refusal, hospital discharge, or death
<sup>b</sup> All PCR-positive NP swabs and air samples, and PCR-positive environmental surface samples with a Ct of $<34.0$ were submitted for virus isolation
<sup>c</sup> Air sampling pumps were calibrated to a flow rate of 3.5 L/min for 2 h; each air sample represents 420 L of air

### NP swabs

A total of 219 follow-up NP swabs were collected. The median number of NP swabs collected per patient was two (IQR 1–4). Overall, 172 (79%) NP swabs from 74 (95%) patients were positive for SARS-CoV-2 by PCR (Table 2). Among patients with at least one positive NP swab, the median number of positive swabs was two (IQR 1–3). The median time between onset of illness and sampling date for PCR-positive swabs was 12 days (IQR 8–17; range 3–52 days).

The median Ct value among positive follow-up NP swabs was 31·4 (IQR 26·8–34·5) (Figure 1). Overall, 30 (27%) of the 110 cultured NP swabs from 21 unique patients yielded viable virus; the highest Ct observed to yield viable virus was 27·2. The median time between onset of illness and sampling date for swabs that yielded viable virus was seven days (IQR 5–11; range 18 days) (Figure 2).

**Figure 2.**
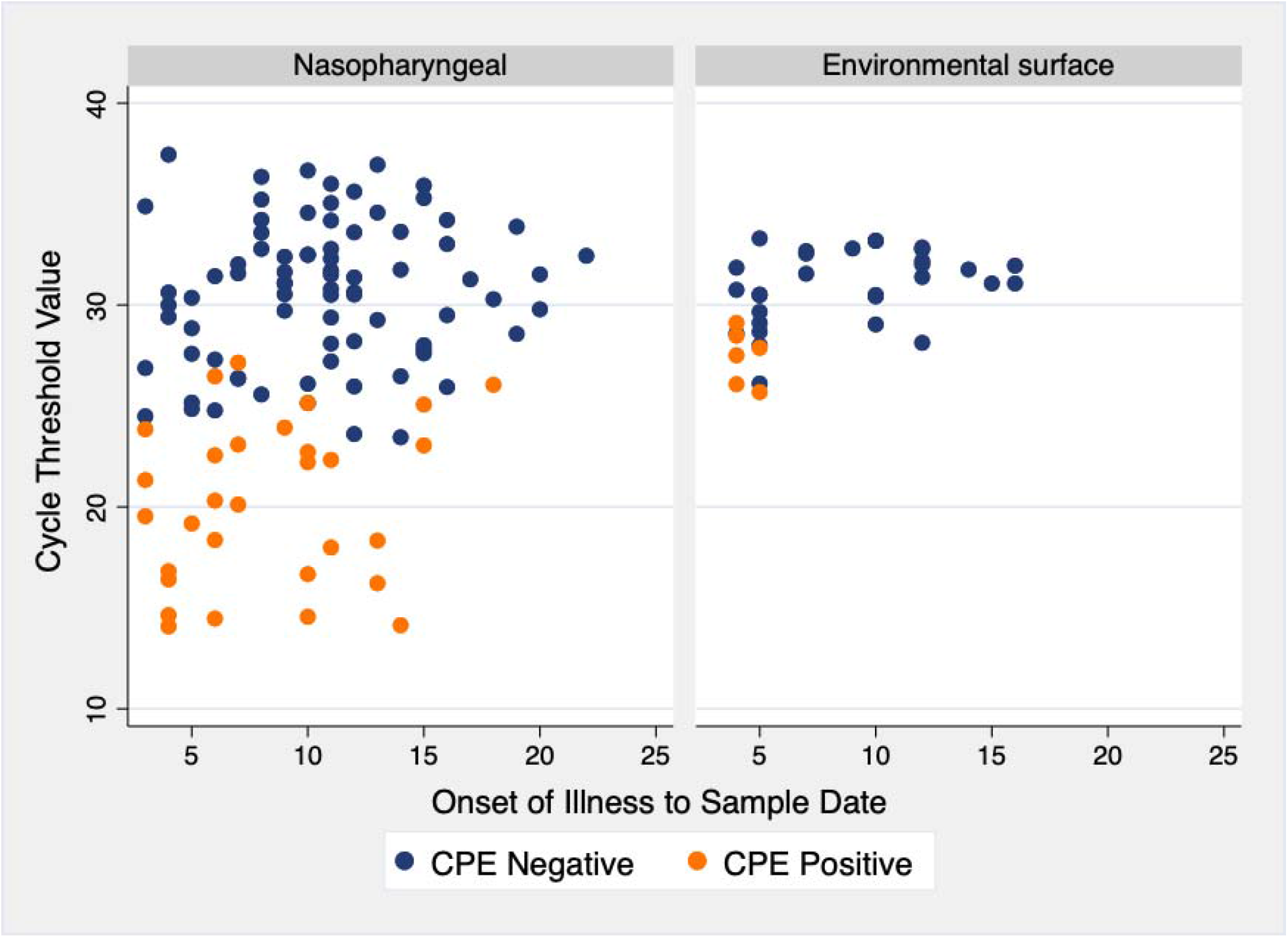
Virus isolation results from 110 nasopharyngeal swabs and 36 surface samples in relation to PCR cycle threshold value and time since symptom onset. CPE, cytopathic effect.

### Surface samples

A total of 474 surface samples were collected. Sixty-one patients (78%) had at least one complete set of surface samples, 12 (15%) had four of five surface types, two (3%) had three, and three (4%) had two. The median duration between onset of illness and surface sample date was ten days (IQR 6–12). The median time between onset of illness and surface sample date for PCR-positive environmental surface samples was nine days (IQR 5–12; range 3–20 days). Overall, 125 (25%) surface samples from 42 (54%) patient rooms yielded SARS-CoV-2 RNA; virus was most frequently detected from the bedrail/ light switch pool and least frequently on the bathroom doorknob (Table 2).

The median Ct value of surface samples testing positive for SARS-CoV2-2 was 33·3 (IQR 29·4–36·0). Cycle thresholds across surface types were similar (Figure 1). Thirty-six surface samples had Ct values <34·0 and six of these (17%), from three (4%) patients, yielded infectious virus; the highest Ct of a sample yielding virus by culture was 29·1 (Figure 2). Viable virus was recovered from each surface type investigated.

### Air samples

A total of 146 air samples were collected; 101 samples (17 gelatin filters, 39 PC filters, 6 PTFE filters, and 13 of each NIOSH stage) at a distance of 1 m from the patient and 45 samples (39 PC filters and 6 PTFE filters) at a distance of 2 m from the patient. Three (2%) air samples taken from three (7%) different rooms at 1 m from the patient were positive for SARS-CoV-2 RNA by PCR; none yielded viable virus. Each of the three PCR-positive air samples were collected by a different air sampling method, including PTFE and PC filters and the NIOSH sampler where viral RNA was detected from stage 1.

### Genome sequencing

In total, 152 surface samples and NP swabs with Ct values ranging from 16·3 to 33·2 (UTR gene) underwent whole genome sequencing for SARS-CoV-2. Fifty passed the CanCOGeN (Canadian COVID Genomics Network) quality control for public release of SARS-CoV-2 genomes and were submitted to GSAID;^24^ 23 of these were from surface samples. Air samples were excluded from the analyses due to poor quality sequences. A phylogenetic analysis of NP and surface swabs is presented in Figure 3. For ease of visualization, we included up to two surface samples per room passing quality control in the phylogenetic analyses. All surface samples cluster with the corresponding NP swabs from patients occupying the same room. The mutations that differentiate the branches on the tree occur in the open reading frames (ORFs) 3, 6, 7 and 8 and genes coding for the nucleocapsid phosphoprotein, the spike protein, and the membrane protein. The N:G321S mutation on the nucleocapsid phosphoprotein was identified from the surface samples and not in the NP swabs in two cases. However, prediction models (SNAP2, PolyPhen-2, SIFT, and MutPred2) of the potential effect on protein functionality showed no predictable gain or loss of function.

**Figure 3.**
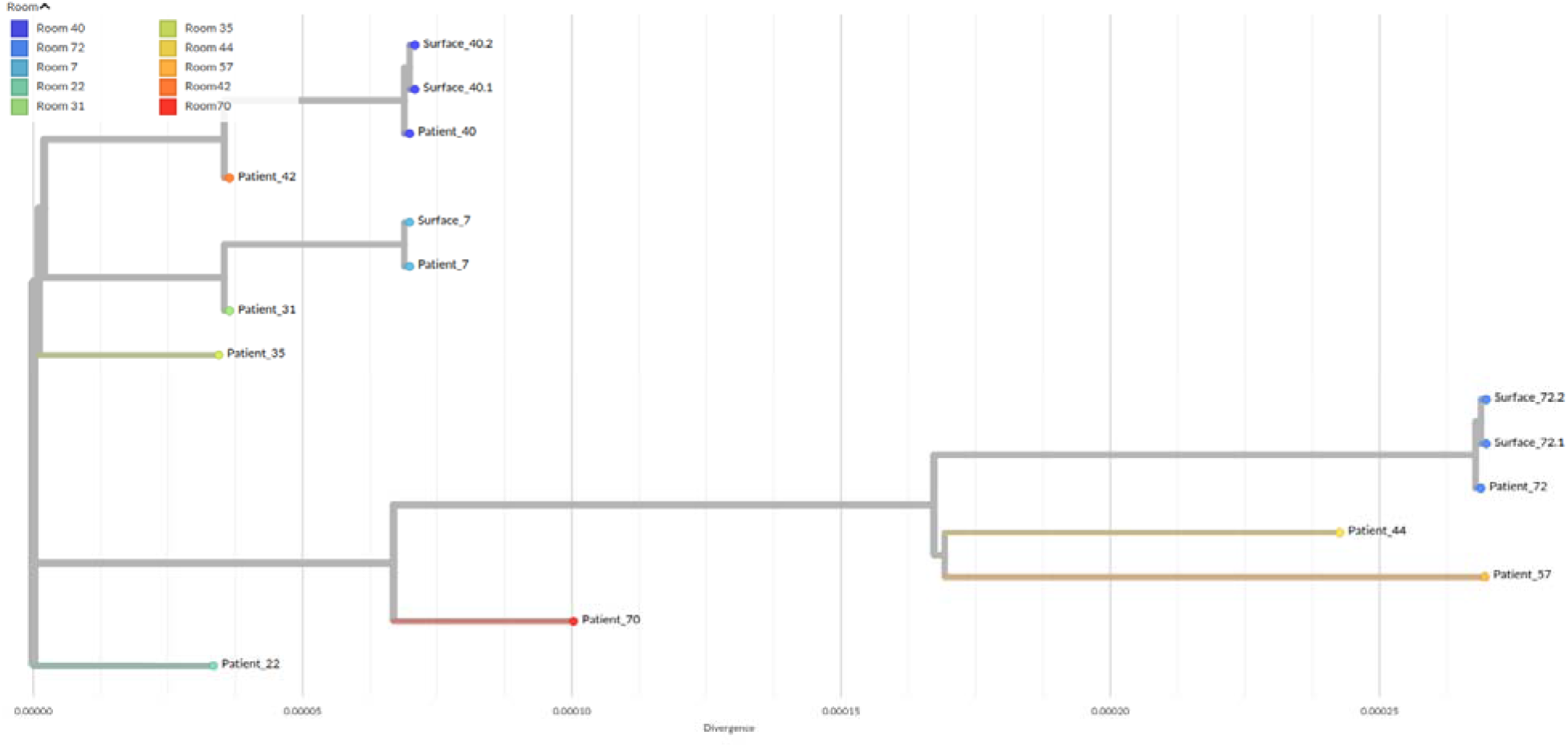
Phylogenetic tree of 15 SARS-CoV-2 genomes from inpatients’ nasopharyngeal swabs and environmental surface swabs from 10 patients’ rooms. Augur pipeline from Nextstain was used to build the phylogenetic tree based on the IQTREE method. The root of the tree is obtained with the first isolate from Wuhan-Hu-1 referenced MN908947·3 in NCBI. The tree is refined using RAxML.

### Factors associated with positive environmental swabs

In the final multivariable mixed-effects model, the following were found to be associated with the detection of SARS-CoV-2 RNA in environmental samples: hypoxia on admission, PCR-positive NP swab with Ct ≤ 30 on or after the environmental sampling date, higher Charlson comorbidity index score, and shorter time from onset of illness to environmental sample date (Table 3).

**Table 3.** Results from the final mixed-effects logistic regression analysis, with a random effect for patient, exploring the association between patient factors and detection of SARS-CoV-2 RNA in environmental samples from 78 hospitalized patients with COVID-19 in Toronto, Canada.

| Explanatory Variable | Category | Odds Ratio (95% CI) | P-Value<sup>a</sup> |
| --- | --- | --- | --- |
| Hypoxic on admission | No | Referent | .. |
|  | Yes | 7.25 (2.00–26.33) | 0.003 |
| PCR-positive nasopharyngeal swab on or after sample date | No | Referent | <0.001 |
|  | Ct >30 | 1.81 (0.30–10.91) | 0.515 |
|  | Ct ≤30 | 15.56 (2.21–109.32) | 0.006 |
| Charlson score | No comorbidities | Referent | 0.006 |
|  | Mild (1-2) | 4.48 (1.28–15.77) | 0.019 |
|  | Moderate–severe (≥3) | 13.72 (2.39–78.80) | 0.003 |
| Onset to sample date | ≤7 days | Referent | .. |
|  | >7 days | 0.24 (0.08–0.72) | 0.011 |
| Variance | Patient | 3.66 (1.73–7.74) | .. |

95% CI, 95% confidence interval.
<sup>a</sup> Significance of variables with three or more categories were assessed via Wald’s chi-square test.

The intraclass correlation coefficient between observations at the patient level was 53% (95% CI: 34– 70%). No outlying observations were identified. Graphical exploration of the BLUPs for patient ID appeared to meet the assumptions of homogeneity of variance and normality.

## Discussion

In this prospective cohort study in Ontario, Canada, SARS-CoV-2 RNA was detected from surfaces (25%) and air (2%) in the acute care setting. Although direct comparison of our results to other studies is limited due to heterogeneity in sampling, processing and detection methodologies, proportionally higher rates of recovery of viral RNA from surfaces compared to the air are broadly consistent with other studies investigating SARS-CoV-2 surface and air contamination.^1,5–7,10–12^

A limited number of studies to date have recovered viable SARS-CoV-2 virus from environmental samples.^14^ We attempted to recover SARS-CoV-2 virus from 36 environmental surface samples, six (17%) of which from three (4%) patients yielded viable virus; positive cultures were confirmed with SARS-CoV-2 RT-PCR. Notably, all surface samples that yielded viable virus were collected from patients within 5 days of illness onset.

PCR-positive air samples were collected from within 1 m of the patient in three cases. However, we were unable to culture viable virus from any of these air samples. To our knowledge only one study has observed CPE from air samples with SARS-CoV-2.^25^ However, it is important to note that the authors concentrated their samples prior to cell culture, potentially optimizing viable virus recovery from samples despite low concentrations of SARS-CoV-2 in the sample. Additionally, although no CPE was observed, Santarpia and colleagues did observe increases in viral RNA in cell culture;^9^ western blot and transmission election microscopy also showed evidence of viral proteins and intact virions. The difficulty in culturing virus from air samples likely relates to a combination of low viral concentrations, dilution effects, the effects of sampling itself on viral cell membrane and surface protein integrity, and duration of air sampling.^3^

The genomic analyses of whole SARS-CoV-2 sequences in the present work confirmed patients were the source of viral contamination of their immediate surroundings in the inpatient setting. There was no clear evidence for a genomic transmission bottleneck in this limited data set.^26^

In the multivariable analysis, hypoxia on admission, a PCR-positive NP swab with a Ct of ≤30 on or after the environmental sampling date, higher Charlson co-morbidity index score, and shorter time from symptom onset to environmental sampling were significantly associated with the detection of SARS-CoV-2 RNA in environmental surface samples (Table 3). Although, to our knowledge, no other study has investigated putative patient factors associated with environmental contamination using multi-variable modelling, our findings are consistent with several observational studies that show that viral load peaks in the first week of illness in COVID-19 patients, with active viral replication in the upper respiratory tract in the first five days of illness.^1,10,27,28^ Additionally, both hypoxia and a high Charlson comorbidity index have previously been found to be associated with higher SARS-CoV-2 viral loads in the nasopharynx.^29,30^

Our study has several limitations. First, although we had a large number of surface and air samples, the samples were recovered from only 78 patients, resulting in a relatively small effective sample size when accounting for clustering. While this still facilitated an exploratory analysis, this limited the power of our multivariable analysis as indicated by the wide confidence intervals for some of the significant variables in our final model. The small effective sample size also prohibited us from investigating factors associated with viable virus in environmental surface samples, including time from symptom onset. Second, the present work focused only on acute care inpatients, excluded critically ill individuals, and had first samples obtained several days after onset of illness. Working in acute care allowed us ready access to patient areas for sampling and clinical data to garner a granular understanding of environmental contamination in hospital settings, the generalizability of our findings to other settings is limited, particularly where room ventilation is highly variable such as homes, schools, long-term care residences, other workplaces, and public spaces. Additionally, it is important to note that these data were collected prior to the emergence of the SARS-CoV-2 variants of concern (VOCs) in late 2020. As such, it is unclear how our results apply to the transmission dynamics of VOCs. Finally, we did not use a standard curve for our RT-PCR analysis and could not calculate the virus concentration per volume of air. Therefore, we were not able to estimate a limit of detection for our aerosol samples.

The findings of this study provide insights into surface and air contamination with SARS-CoV-2 in hospitalized COVID-19 patients. We found that SARS-CoV-2 RNA was detected on a minority of surfaces in COVID-19 patients’ rooms and rarely from air samples, suggesting these sources are unlikely to pose a major exposure risk in hospitals with similar surface decontamination procedures and ventilation in place. Additionally, hypoxia on admission, PCR-positive NP swab on or after sampling date, co-morbidities, and time since symptom onset were found to be important factors associated with the detection of SARS-CoV-2 RNA in the environment. These findings suggest that while early detection and isolation of COVID-19 patients are important, air and surfaces may pose limited risk a few days after admission to acute care hospitals.

## Data Availability

The data that support the findings of this study are available from the corresponding author, SM, upon reasonable request.

## Contributors

This study was conceived, designed, and supervised by AJM and SM. Air sampling protocols were designed by SM, MV, CD, KP, and HM. Sample collection and laboratory analysis was carried out by SB, NG, GC, AF, LF, SK, KN, AXI, HPM, MM, AP, SP, KP, RS, LY, PA, and XZZ. AF, LF, SK, and RH conducted patient chart reviews. Genome sequencing and data analysis were done by AJJ, JDK, HM, JAN, EMP, and AGM. JDK, AJJ, HM, AJM, AGM and SM interpreted the data. JDK and AJJ had full access to all the data in the study. The manuscript was drafted by JDK and AJJ and was critically revised by all authors.

## Declarations of interest

All authors report no conflicts relevant to this article.

## Acknowledgments

This study was supported by the Canadian Institutes of Health Research (CIHR No. RN419944-439999) as well as Genome Canada CanCOGeN funding. Computational support was provided by the McMaster Service Lab and Repository (MSLR) computing cluster, supplemented by hardware donations and loans from Cisco Systems Canada, Hewlett Packard Enterprise, and Pure Storage. Many thanks to the patients with COVID-19 who agreed to participate in this research. We are grateful to the laboratory, infection prevention and control, public health, and research ethics staff who make the Toronto Invasive Bacterial Diseases Network possible. JDK is supported by an AMMI Canada/BioMérieux Fellowship in Microbial Diagnostics. AJJ was supported by the Vanier Canada Graduate Scholarship at the time of this work. HM is funded with a postdoctoral fellowship from le Fond de Recherche du Québec – Nature et Technologie and is the recipient of the Lab Exchange Visitor Program Award from the Canadian Society for Virology. AGM holds McMaster’s inaugural David Braley Chair in Computational Biology, generously supported by the family of the late Mr. David Braley.

## Notes

### Competing Interest Statement

The authors have declared no competing interest.

### Author Declarations

Research ethics approval was granted by all participating The Toronto Invasive Bacterial Diseases Network hospitals (Sunnybrooks Research Ethics Board, The Mount Sinai Hospital Research Ethics Board, Toronto East Health Network Research Ethics Board, Oslers Research Ethics Board, and Scarborough Health Networks Research Ethics Board).

### Summary of Updates

A mistake was found in the denominator of surface samples investigated via viral culture. Thirty-six samples with a ct of less than 34.0 were assessed, not 42. Any mention of this throughout the text was updated. No other changes to the manuscript or supplemental materials were made.

