## Supplemental Material for "Surface and air contamination with SARS-CoV-2 from hospitalized COVID-19 patients in Toronto, Canada"

**Supplementary Methods**

*Genome Sequencing*

The ARTIC V3 SARS-CoV-2 protocol (<https://artic.network/ncov-2019>) was used for amplicon generation. Briefly, 1 µL Random Primer Mix (ProtoScript II First Strand cDNA Synthesis Kit, New England Biolabs, NEB, USA) and 1 µL 10mM dNTP mix (NEB, USA) was added to 8 µL extracted RNA and denatured at 65 °C for 5 min and then incubated on ice. 12·5 µL 2X ProtoScript II Reaction Mix and 2·5 µL 10X ProtoScript II Enzyme Mix were then added to the denatured sample and cDNA synthesis performed using the following conditions: 25 °C for 5 min, 42 °C for 50 min and 80 °C for 5 min.

After cDNA synthesis two multiplex PCR tiling reactions were prepared. In a PCR tube, 2·5 µL cDNA was combined with 12·5 µL Q5 High-Fidelity 2X Master Mix (NEB, USA). To mix #1 5·87 µL nuclease free water (Thermo Fisher Scientific, USA), and 4·13 µL of 10 µM ARTIC v3 primer pool #1 was added. To mix #2 5·95 µL nuclease free water and 4·05 µL of 10 µM ARTIC v3 primer pool #2 was added. PCR cycling was then performed as follows: 98 °C for 30 s followed by 35 cycles of 98 °C for 15 s and 65 °C for 5 min.

PCR products were combined, and a clean-up was performed with AMPure XP beads (Beckman Coulter, USA) at a 1:1 bead to sample ratio. The quantity of amplicons was measured with the Qubit 4·0 fluorometer using the 1X dsDNA HS Assay Kit (Thermo Fisher Scientific, USA). The sequencing libraries were prepared using the Nextera DNA Flex Prep kit (Illumina, USA) as per manufacturer’s instructions.

Paired-end (2x150 bp) sequencing was performed on a MiniSeq with a 300–cycle reagent kit (Illumina, USA). A negative control library with no input SARS-CoV-2 RNA extract was included in the sequencing run.

*Genome assembly and sequence analyses*

Short-read sequence processing and genome assembly was done using the SIGNAL (SARS-CoV-2 Illumina GeNome Assembly Line) pipeline (<https://github.com/jaleezyy/covid-19-signal>; (Nasir et al., 2020). Within SIGNAL, the consensus genome sequences are generated based on the SARS-CoV-2 reference genome (MN908947.3 NCBI reference – i.e. the first isolate from Wuhan-Hu-1) using iVAR consensus generation. MAFFT (Multiple alignment using fast Fourier transform) was used for multiple sequence alignment of the consensus sequences. Nextstrain with the augur pipeline was used to build a maximum liklihood tree based on the IQTREE method. The tree is refined using RAxML (Randomized Axelerated Maiximum Likelihood). Auspice was then used to visualize and interact with the tree.

**Supplementary Table 1.** Number and types of samples collected from 78 COVID-19 inpatients in Toronto, Canada

| **Room ID** | **Air samples** | | | **Environmental samples** | | | | | | **NP swabs** | **Grand Total** |
| --- | --- | --- | --- | --- | --- | --- | --- | --- | --- | --- | --- |
|  | **1 m** | **2 m** | **Total** | **Bathroom door** | **Bed and switch** | **Phone** | **Table and chair** | **Toilet and sink** | **Total** |  |  |
| 1 | 1 | 1 | 2 | 2 | 2 | 2 | 2 | 2 | 10 | 2 | 14 |
| 2 |  |  |  | 1 | 1 | 1 | 1 | 1 | 5 | 3 | 8 |
| 3 |  |  |  | 1 | 1 | 1 | 1 | 1 | 5 | 2 | 7 |
| 4 | 1 | 1 | 2 | 1 | 1 | 1 | 1 | 1 | 5 | 2 | 9 |
| 5 |  |  |  |  | 2 | 1 | 2 | 2 | 7 | 6 | 13 |
| 6 |  |  |  | 1 | 2 | 1 | 1 | 3 | 8 | 5 | 13 |
| 7 | 1 | 1 | 2 | 1 | 1 | 1 | 1 | 1 | 5 | 1 | 8 |
| 8 |  |  |  | 1 | 1 | 1 | 1 | 1 | 5 | 1 | 6 |
| 9 |  |  |  | 1 | 1 | 1 | 1 | 1 | 5 | 1 | 6 |
| 10 | 1 | 1 | 2 | 1 | 1 | 1 | 1 | 1 | 5 | 2 | 9 |
| 11 | 1 | 1 | 2 |  | 1 | 1 | 1 | 2 | 5 | 2 | 9 |
| 12 | 1 | 1 | 2 | 1 | 1 | 1 | 1 | 1 | 5 | 1 | 8 |
| 13 |  |  |  | 1 | 1 | 1 | 1 | 1 | 5 | 2 | 7 |
| 14 |  |  |  | 1 | 1 | 1 | 1 | 1 | 5 | 1 | 6 |
| 15 | 1 | 1 | 2 | 2 | 2 | 1 | 2 | 2 | 9 | 3 | 14 |
| 16 |  |  |  | 1 | 1 | 1 | 1 | 1 | 5 | 4 | 9 |
| 17 | 1 | 1 | 2 |  | 1 | 1 | 1 | 1 | 4 | 3 | 9 |
| 18 |  |  |  |  | 1 |  | 1 |  | 2 | 10 | 12 |
| 19 |  |  |  | 1 | 1 | 1 | 1 | 1 | 5 | 2 | 7 |
| 20 |  |  |  |  | 1 | 1 |  |  | 2 | 1 | 3 |
| 21 | 1 | 1 | 2 | 2 | 2 | 2 | 2 | 2 | 10 | 4 | 16 |
| 22 |  |  |  |  | 1 |  | 1 | 1 | 3 | 12 | 15 |
| 23 |  |  |  | 2 | 2 | 2 | 2 | 2 | 10 | 3 | 13 |
| 24 |  |  |  | 1 | 1 | 1 | 1 | 1 | 5 | 1 | 6 |
| 25 | 1 | 1 | 2 | 2 | 2 | 2 | 2 | 2 | 10 | 2 | 14 |
| 26 |  |  |  | 1 | 1 | 1 | 1 | 1 | 5 | 1 | 6 |
| 27 |  |  |  | 2 | 2 |  | 2 | 2 | 8 | 4 | 12 |
| 28 | 2 | 1 | 3 | 1 | 1 | 1 | 1 | 1 | 5 | 2 | 10 |
| 29 |  |  |  | 1 | 1 | 1 | 1 | 1 | 5 | 1 | 6 |
| 30 | 1 | 1 | 2 | 1 | 1 | 1 | 1 | 1 | 5 | 2 | 9 |
| 31 | 1 | 1 | 2 | 1 | 1 | 2 |  | 1 | 5 | 1 | 8 |
| 32 |  |  |  | 1 | 1 | 1 |  | 2 | 5 | 2 | 7 |
| 33 |  |  |  | 1 | 1 | 1 | 1 | 1 | 5 | 2 | 7 |
| 34 |  |  |  | 1 | 1 |  | 1 | 1 | 4 | 2 | 6 |
| 35 |  |  |  | 1 | 1 |  | 1 | 1 | 4 | 1 | 5 |
| 36 |  |  |  | 1 | 1 | 1 | 1 | 1 | 5 | 1 | 6 |
| 37 | 2 | 1 | 3 | 1 | 1 | 1 | 1 | 1 | 5 | 1 | 9 |
| 38 | 2 | 1 | 3 | 2 | 2 | 2 | 1 | 3 | 10 | 7 | 20 |
| 39 | 2 | 1 | 3 | 1 | 1 | 1 | 1 | 1 | 5 | 4 | 12 |
| 40 |  |  |  | 3 | 4 | 3 | 4 | 4 | 18 | 6 | 24 |
| 41 | 2 | 1 | 3 | 2 | 2 | 2 | 2 | 2 | 10 | 4 | 17 |
| 42 | 2 | 1 | 3 | 1 | 1 | 1 | 1 | 1 | 5 | 1 | 9 |
| 43 | 2 | 1 | 3 | 1 | 1 | 1 | 1 | 1 | 5 | 2 | 10 |
| 44 |  |  |  |  | 1 |  | 1 | 1 | 3 | 13 | 16 |
| 45 |  |  |  | 1 | 1 | 1 | 1 | 1 | 5 | 1 | 6 |
| 46 | 2 | 1 | 3 |  | 1 | 1 | 1 | 1 | 4 | 1 | 8 |
| 47 |  |  |  |  | 1 |  | 1 |  | 2 | 5 | 7 |
| 48 |  |  |  | 2 | 2 | 2 | 2 | 2 | 10 | 4 | 14 |
| 49 |  |  |  | 1 | 1 | 1 | 1 | 1 | 5 | 1 | 6 |
| 50 |  |  |  | 1 | 2 | 1 |  | 2 | 6 | 3 | 9 |
| 51 | 2 | 1 | 3 | 2 | 2 | 2 | 2 | 2 | 10 | 4 | 17 |
| 52 |  |  |  | 1 | 2 | 1 | 2 |  | 6 | 3 | 9 |
| 53 | 2 | 1 | 3 | 1 | 1 | 1 | 1 | 1 | 5 | 2 | 10 |
| 54 |  |  |  | 1 | 1 | 1 | 1 | 1 | 5 | 1 | 6 |
| 55 | 2 | 1 | 3 | 1 | 1 | 1 | 1 | 1 | 5 | 1 | 9 |
| 56 |  |  |  | 1 | 1 |  | 1 | 1 | 4 | 1 | 5 |
| 57 | 2 | 1 | 3 | 1 | 1 | 1 | 1 | 1 | 5 | 2 | 10 |
| 58 | 2 | 1 | 3 | 1 | 1 | 1 | 1 | 1 | 5 | 2 | 10 |
| 59 | 2 | 1 | 3 | 1 | 1 | 1 | 1 | 1 | 5 | 1 | 9 |
| 60 | 2 | 1 | 3 | 2 | 2 | 2 | 2 | 2 | 10 | 4 | 17 |
| 61 | 2 | 1 | 3 | 1 | 1 | 1 | 1 | 1 | 5 | 3 | 11 |
| 62 | 2 | 1 | 3 | 1 | 1 | 1 | 1 | 1 | 5 | 1 | 9 |
| 63 | 4 | 1 | 5 | 1 | 1 | 1 | 1 | 1 | 5 | 9 | 19 |
| 64 | 4 | 1 | 5 | 2 | 2 | 2 | 2 | 2 | 10 | 3 | 18 |
| 65 | 4 | 1 | 5 | 1 | 1 | 1 | 1 | 1 | 5 | 1 | 11 |
| 66 | 4 | 1 | 5 | 1 | 1 | 1 | 1 | 1 | 5 | 8 | 18 |
| 67 | 1 | 1 | 2 | 1 | 1 | 1 | 1 | 1 | 5 | 1 | 8 |
| 68 | 1 | 1 | 2 | 1 | 1 | 1 | 1 | 1 | 5 | 1 | 8 |
| 69 | 4 | 1 | 5 | 2 | 2 | 2 | 2 | 2 | 10 | 1 | 16 |
| 70 | 1 | 1 | 2 | 1 | 1 | 1 | 1 | 1 | 5 | 2 | 9 |
| 71 | 4 | 1 | 5 | 2 | 2 | 2 | 2 | 2 | 10 | 2 | 17 |
| 72 | 4 | 1 | 5 | 3 | 3 | 3 | 3 | 3 | 15 | 7 | 27 |
| 73 | 4 | 1 | 5 | 1 | 1 | 1 | 1 | 1 | 5 | 1 | 11 |
| 74 | 4 | 1 | 5 | 1 | 1 | 1 | 1 | 1 | 5 | 1 | 11 |
| 75 | 4 | 1 | 5 | 1 | 1 | 1 | 1 | 1 | 5 | 2 | 12 |
| 76 | 4 | 1 | 5 | 1 | 1 | 1 | 1 | 1 | 5 | 4 | 14 |
| 77 | 4 | 1 | 5 | 2 | 2 | 2 | 2 | 2 | 10 | 2 | 17 |
| 78 | 4 | 1 | 5 | 1 | 1 | 1 | 1 | 1 | 5 | 1 | 11 |
| Grand Total | 101 | 45 | 146 | 88 | 102 | 88 | 95 | 101 | 474 | 219 | 839 |

**Supplementary Table 2.** Patient demographics and clinical characteristics for 474 environmental surface samples collected from 78 COVID-19 inpatients in Toronto, Canada

| **Patient Characteristics** | **No. patients (%)**  **(N=474)** |
| --- | --- |
| Age: <65 years  ≥65 years | 191 (40)  283 (60) |
| Sex (number (%) male) | 267 (56) |
| Charlson comorbidity index: 0  1-2  ≥3 | 205 (43)  190 (40)  79 (17) |
| Underlying chronic illness: Diabetes mellitus  Pulmonary  Cardiac | 136 (29)  91 (19)  10 (22) |
| History of smoking | 133 (28) |
| Clinical Frailty Score (n=470)^a^: Not frail (1-4)  Mild to moderate (5-6)  Severe (7-8) | 314 (67)  106 (22)  50 (11) |
| Symptoms/signs: Cough  Fever  Diarrhea  Delirium/confusion  O_2_ saturation <92% at admission | 385 (81)  370 (78)  150 (32)  78 (16)  248 (52) |
| Oxygen requirements during admission:  No oxygen required  Required oxygen by face mask or nasal prong only  Required high flow oxygen  Required intubation | 125 (26)  273 (58)  57 (12)  67 (14.14) |
| Required oxygen by facemask/nasal prong | 337 (71) |
| Required high flow oxygen, not intubated | 38 (8) |
| Management Prone positioning  Received steroids (day of sampling)  ICU admission (day of sampling) | 34 (7)  30 (6)  91 (19) |
| Accommodation: Regular private room  Negative pressure room | 195 (41)  279 (59) |
| Onset of illness to sample date: ≤7 days  >7 days | 150 (32)  324 (68) |

**^a^** The clinical frailty was collapsed into three categories: non-frail (1–4), mild-to-moderately frail (5–6), and severely frail (7–8)

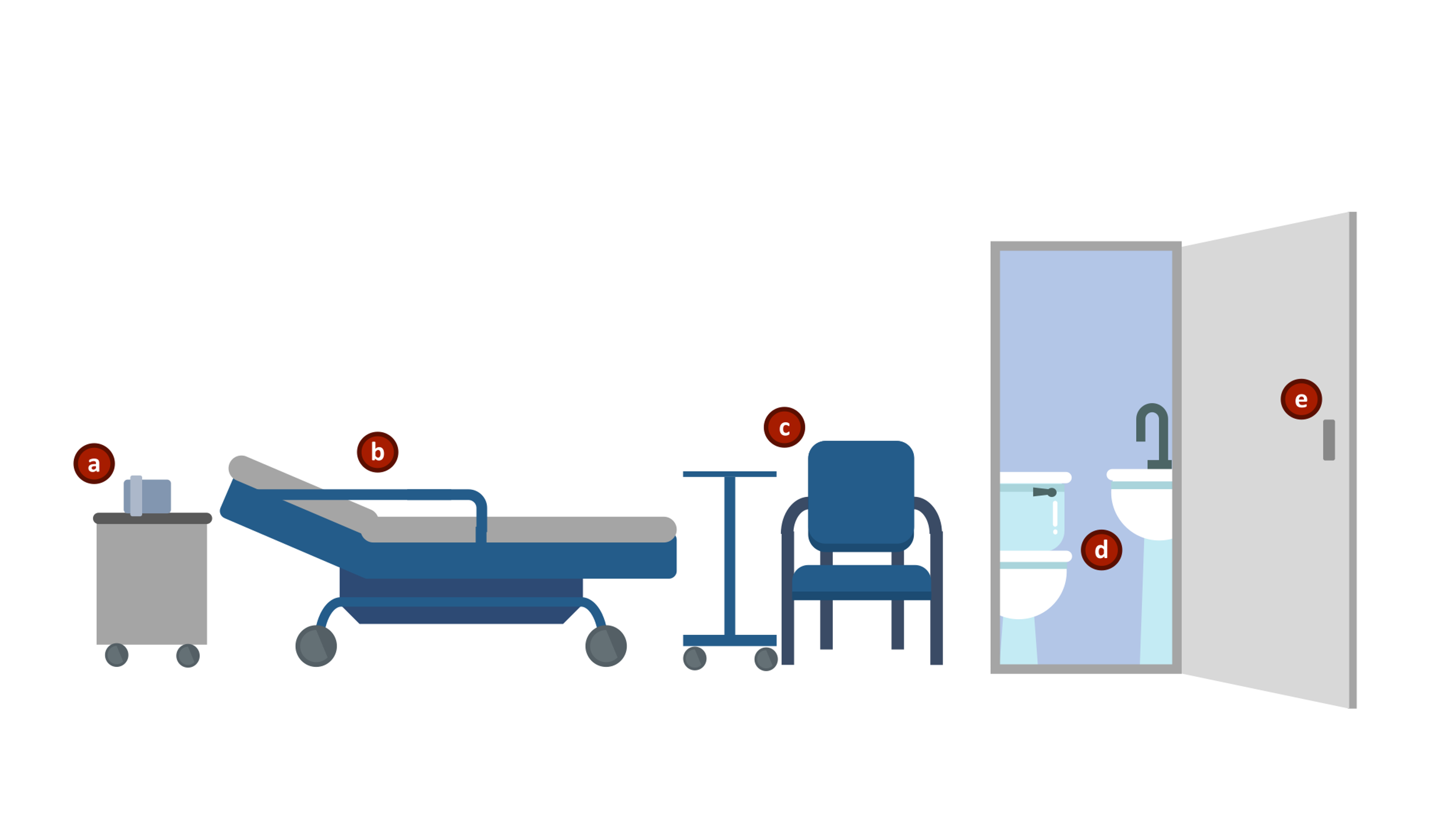

**Supplementary Figure 1**. Environmental surface sampling. Surface samples were collected from the following surfaces: a) phone (all surfaces of the patient’s phone and room phone), b) bed (bed rail and pillow) and light switch or pullcord in patient’s bedspace (pooled), c) overbed table and chair (pooled), d) toilet and sink faucet handles (pooled), and e) bathroom doorknob.
